# Diagnostic Accuracy of Artificial Intelligence for Arrhythmia Detection Using the 12-Lead Electrocardiogram: A Systematic Review and Meta-Analysis

**DOI:** 10.64898/2026.02.06.26345251

**Authors:** Luiz Filipe Torres de Alencar, Gabriel Fontenele Ximenes, Marina de Andrade Norões Bezerra, Leonardo Brito de Souza, Nicole Aires Perazolo, João Pedro Teixeira Bentes Monteiro, Pedro Jorge Pires Viana, Mateus Paiva Marques Feitosa, Jefferson Luís Vieira, Shaan Khurshid

## Abstract

**Background:** Artificial intelligence (AI) has emerged as a promising tool for interpreting 12-lead electrocardiograms (ECGs), with the potential to enhance diagnostic accuracy for arrhythmia detection. However, published studies vary widely in methodology and validation strategy, warranting a quantitative synthesis of diagnostic performance.

**Methods:** A systematic review and meta-analysis was conducted according to the PRISMA-DTA 2018 guidelines and registered in PROSPERO (CRD420251027264). Searches were performed in MEDLINE, Embase, and Cochrane Library through September 2025 without language restrictions. Studies evaluating AI algorithms for arrhythmia detection using 12-lead ECGs were included. Data on sensitivity, specificity, and area under the curve (AUC) were extracted. Pooled estimates were generated using a bivariate random-effects model. Risk of bias was assessed with QUADAS-2, and the certainty of evidence was quantified using GRADE.

**Results:** 20 studies were included in the meta-analysis, encompassing over 5.5 million ECGs. The pooled sensitivity, specificity, and AUC for AI-based arrhythmia detection were 94.0% (95% CI 90.8–96.2; I² = 96.9%), 98.7% (95% CI 97.3–99.3; I² = 98.3%), and 0.982 (95% CI 0.965–0.986), respectively. Detection of atrial fibrillation (AF) yielded a sensitivity of 92.6% (95% CI 86.4–96), a specificity of 99.1% (95% CI 98.4–99.5), and an AUC of 0.988. Convolutional neural networks (CNN) specifically demonstrated a sensitivity of 97.6%, specificity of 98.7%, and an AUC of 0.982 for overall arrhythmia detection. When limited to external validation (n=6), the sensitivity was 96.9% (95% CI 89.2–99.1), specificity was 95.6% (95% CI 77.6–99.3), and AUC was 0.983. No significant publication bias was detected, and the overall certainty of evidence was rated as high.

**Conclusions:** AI models applied to 12-lead ECGs demonstrate excellent diagnostic performance for arrhythmia detection. Findings support potential integration into clinical workflows, particularly in settings with limited cardiology expertise. Given substantial heterogeneity, standardized datasets and multicenter prospective validation are essential to ensure effective and equitable implementation.

**What is Known:**

- Artificial intelligence has been increasingly applied to 12-lead electrocardiograms for arrhythmia detection, with multiple studies reporting high diagnostic accuracy.

**What the Study Adds:**

- This meta-analysis demonstrates consistently high diagnostic performance of artificial intelligence for arrhythmia detection on 12-lead ECGs, including atrial fibrillation and externally validated models.
- The substantial heterogeneity observed underscores the need for standardized datasets and multicenter prospective validation before widespread clinical implementation.

## Introduction

Cardiac arrhythmias comprise abnormalities in cardiac rhythm and represent a heterogeneous group of conditions ^1^. Clinically, they range from largely benign conditions (e.g., isolated premature contractions) to highly morbid arrhythmias (e.g., atrial fibrillation [AF]) and even conditions capable of causing sudden death (e.g., ventricular tachycardia and ventricular fibrillation). Their timely and accurate identification is essential, as prompt management may reduce the substantial morbidity and mortality burden imposed by arrhythmias worldwide ^2^.

The 12-lead electrocardiogram (ECG) stands as a cornerstone in the diagnosis and monitoring of cardiac disorders, providing a noninvasive window into the heart’s electrical activity ^3^. Despite standardized acquisition protocols, manual interpretation requires substantial expertise and is prone to interobserver variability ^4^. Consequently, the need for more consistent and efficient ECG analysis methods has driven the integration of computational tools into routine clinical workflows. Advances in computational power, combined with the exponential expansion of available ECG datasets, are providing new opportunities for innovation in ECG interpretation ^5^.

Recent advances in artificial intelligence (AI) have revolutionized ECG interpretation ^6^. AI models can identify subtle nonlinear patterns within ECG signals, and have been reported to achieve ECG interpretation accuracy comparable to or even exceeding that of cardiologists ^7^. Indeed, some studies have reported accuracies above 98% for arrhythmia classification and strong predictive performance in detecting atrial fibrillation ^8,9^. However, many of these studies rely on small datasets without external validation, limiting confidence in the precision and potential generalizability of reported metrics. Therefore, we conducted a systematic review and meta-analysis to quantitatively synthesize diagnostic accuracy measures (sensitivity, specificity, and area under the receiver operating characteristic curve (AUC)) of AI models using 12-lead ECGs for arrhythmia detection.

## Methods

### Overview

This study followed the guidelines of the Preferred Reporting Items for Systematic Reviews and Meta-Analysis for Diagnostic Test Accuracy (PRISMA-DTA) 2018 protocol^10^. The last search was performed in September 2025, using the MEDLINE (via PubMed), Embase, and Cochrane Library databases. Databases were searched using the appropriate descriptors for each platform without considering a specific language or time frame. The descriptors “Arrhythmias, Cardiac”[MeSH], “Arrhythmia”, “Cardiac Arrhythmia”, “Atrial Fibrillation”, “Atrial Flutter” OR “Supraventricular Tachycardia”, “Ventricular Tachycardia”, “Bradyarrhythmia” AND “Artificial Intelligence”[MeSH], “Machine Learning”, OR “Deep Learning”[MeSH], “Neural Networks, Computer”[MeSH], “Electrocardiography”[MeSH] OR “Electrocardiogram” OR “ECG” OR “EKG” were used and combined utilizing the Boolean operators AND and OR. The references of all included studies were also reviewed for additional publications. The study protocol was registered on the PROSPERO platform under the protocol CRD420251027264.

### Eligibility Criteria

The PRISMA flow diagram is available in **Figure 1**. Initially, 3813 articles were identified and screened, with independent analysis by two researchers (NAP; PJV), and disagreements resolved by consensus with a third reviewer (LFA). After excluding duplicates, 2991 studies remained, and their titles and abstracts were judged based on the PICOS model for inclusion criteria as follows: (1) participants (patients who underwent ECG recording); (2) intervention (arrhythmia diagnosis by artificial intelligence on ECG); (3) control (none); (4) outcome (sensitivity, specificity, and area under the curve [AUC]); and (5) study design (diagnostic studies). The exclusion criteria were: (1) studies employing AI models unrelated to diagnostic image or signal processing (e.g., language models or natural language processing-based algorithms); (2) studies that did not report sensitivity, specificity, AUC, or sufficient data to calculate these indicators; and (3) reports other than original research (e.g., case reports, editorials, letters to the editor, and reviews).

**Fig. 1.**
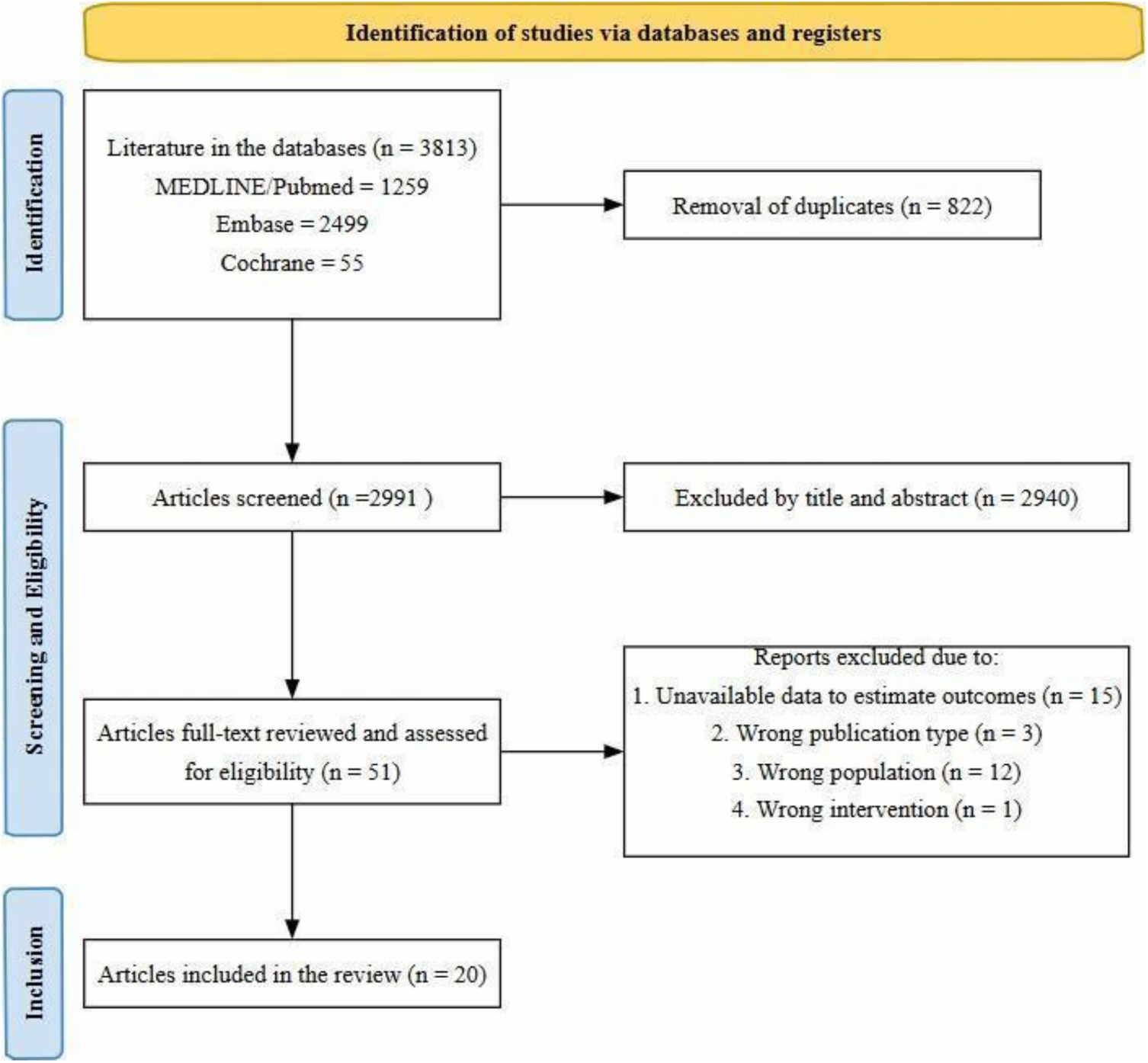
Flowchart of article selection according to PRISMA 2020.

### Quality Assessment

Two study authors (JPM; LBS) independently assessed the risk of bias using the Quality Assessment of Diagnostic Accuracy Studies (QUADAS-2) tool ^11^. Discordances were resolved by consensus. The analysis includes four main domains, including i) patient selection, ii) index test, iii) reference standard, and iv) how patient flow occurred and the timing of the index test and reference standard (flow and timing). All four domains were assessed for risk of bias, and the first three were also evaluated for concerns about applicability. Each domain could be answered with yes, no, or unclear. In addition, the tool consisted of four phases: reporting the review question, providing review-specific guidance, reviewing or developing the flow diagram, and finally, judging bias and applicability.

We used the Grading of Recommendations Assessment, Development, and Evaluation (GRADE) assessment tool ^12^ to rate the certainty of the evidence, as implemented in the GRADE Pro version 3.6 software (McMaster University).

To assess publication bias, we performed funnel plot analysis of point estimates by study weight. We performed an Egger regression test to assess funnel plot asymmetry and potential publication bias, as more than 10 studies were included in the meta-analysis. A p-value < 0.05 was considered indicative of significant asymmetry.

### Data Extraction

Two authors (NAP; MAN) independently extracted the data from all the eligible studies. To ensure the integrity and accuracy of the information, discordances were discussed and resolved by consensus with a third reviewer (LFA). We extracted the following data from the eligible studies: first author’s last name, year and country of publication, study design, study period, total number of patients, total number of ECGs, number of ECGs with and without arrhythmias in the test set, percentage of male patients, mean age, classes of arrhythmias in the study, AI architecture, standard of reference for diagnosis, accuracy, F1 score, and data necessary to calculate sensitivity, specificity, and AUC. If it was not possible to calculate the outcomes for all arrhythmias in the study, we extracted the data for AF, which was the most common target label.

### Outcomes

Our primary study goal was to assess the overall diagnostic performance of AI models applied to 12-lead ECGs for the detection of cardiac arrhythmias, quantified as sensitivity, specificity, and area under the receiver operating characteristic curve (AUC). We extracted rates of true positives, false positives, true negatives, and false negatives, and conducted a bivariate random-effects meta-analysis to generate pooled estimates accounting for the inherent correlation between sensitivity and specificity.

In addition to overall diagnostic performance, we sought to: (1) evaluate the clinical robustness of AI-ECG models when subjected to external validation, (2) quantify the potential effects of model architecture, (3) analyze the effectiveness of these models in the detection of a single arrhythmia class (AF) and (4) assess the impact of the prevalence of positives cases in the data heterogeneity. Therefore, we performed secondary analyses in which we fit analogous bivariate models within studies grouped by validation framework, algorithmic approach, AF and prevalence of abnormal classes.

### Data Analysis

Statistical analysis was performed according to the guidelines of the Cochrane Collaboration and the PRISMA 2020 statement ^14^. We performed all meta-analyses considering the best model’s performance in each study’s test set using the random-effects model approach. The random-effects model was preferred as it is more robust to heterogeneity in effects across studies, which was anticipated due to substantial differences across machine learning models and the common lack of standardization across AI studies (e.g., patient characteristics, label definitions, feature selection, and outcome measurement). Proportional analysis with logit transformation was used for univariate analysis. For bivariate analysis, a summary receiver operating characteristic curve (SROC) was constructed. Study heterogeneity was estimated using I², τ², and the Cochran Q test, with I² > 50% considered indicative of meaningful heterogeneity. To explore heterogeneity, we performed subgroup analysis, sensitivity analysis, and meta-regression with continuous variables. All statistical tests were two-tailed. Statistical analysis was performed with R software version 4.4.2.

## Results

### Characteristics of Included Studies

After the screening and eligibility processes, 20 retrospective cohorts were included ^13–33^. **Table 1** summarizes the baseline characteristics of the studies. Investigations were conducted across multiple regions, including North and South America, Europe, Asia, Canada, Egypt, Russia, the United States, China, Bulgaria, South Korea, Taiwan, Israel, and Brazil. Six studies performed external validation; one ^24^ was included only in the external validation subgroup analysis, as it was not possible to estimate the internal validation data. Included studies encompassed 5,521,809 ECGs (including development, training, and test sets) and 1,935,368 patients, with an average age of 58.9 years and a mean male proportion of 53.5%. The test sets contained 92,159 ECGs, of which 66,031 (71.7%) were sinus rhythm, and 20,015 (21.7%) represented arrhythmias. All studies analyzed standard 12-lead ECGs for arrhythmia detection or classification. Some focused on a single rhythm, most commonly AF, whereas others examined a broader arrhythmic spectrum, including atrial flutter, bundle branch blocks, premature complexes, supraventricular tachycardia, and ventricular tachycardia.

**Table 1:**
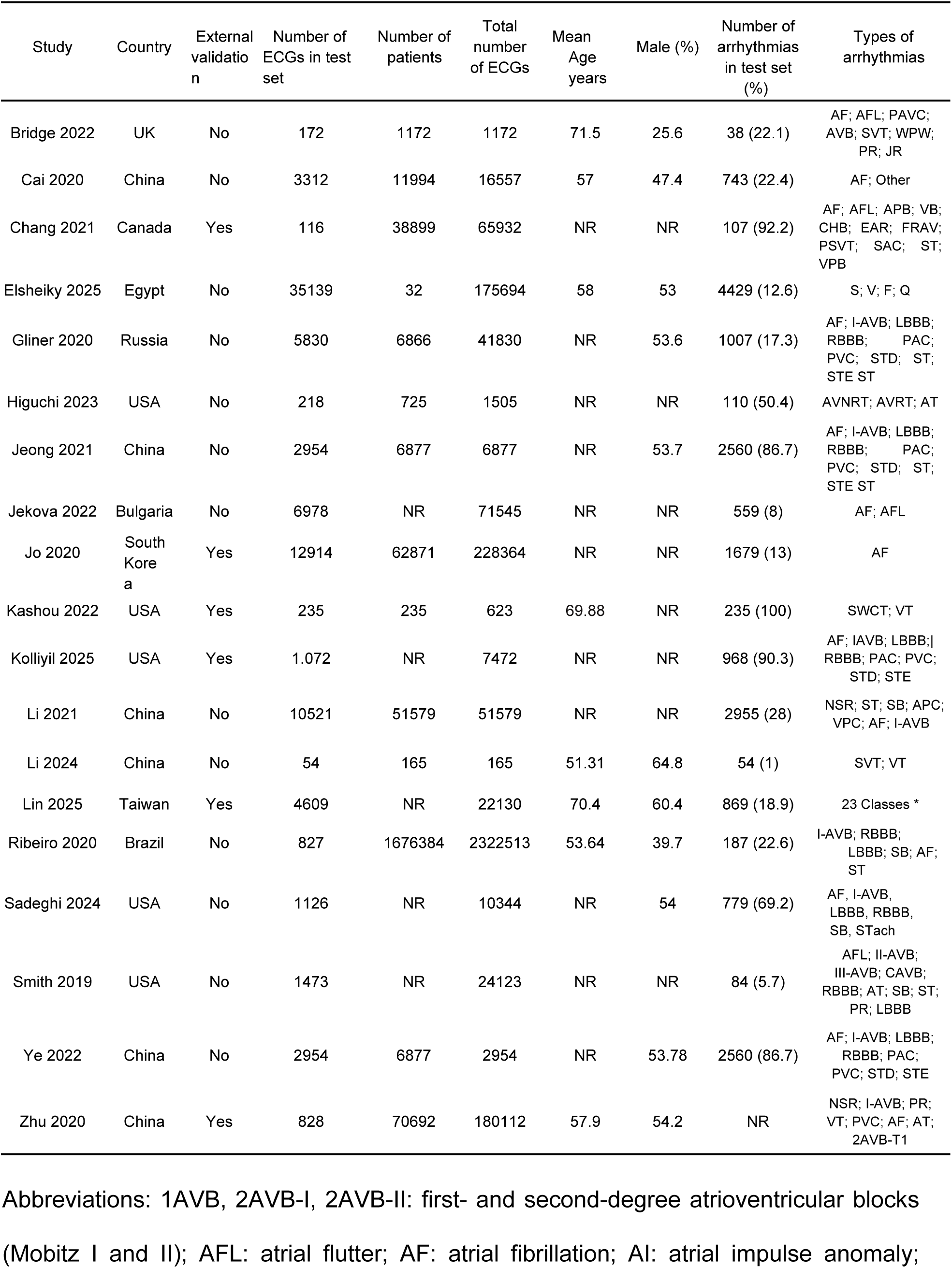
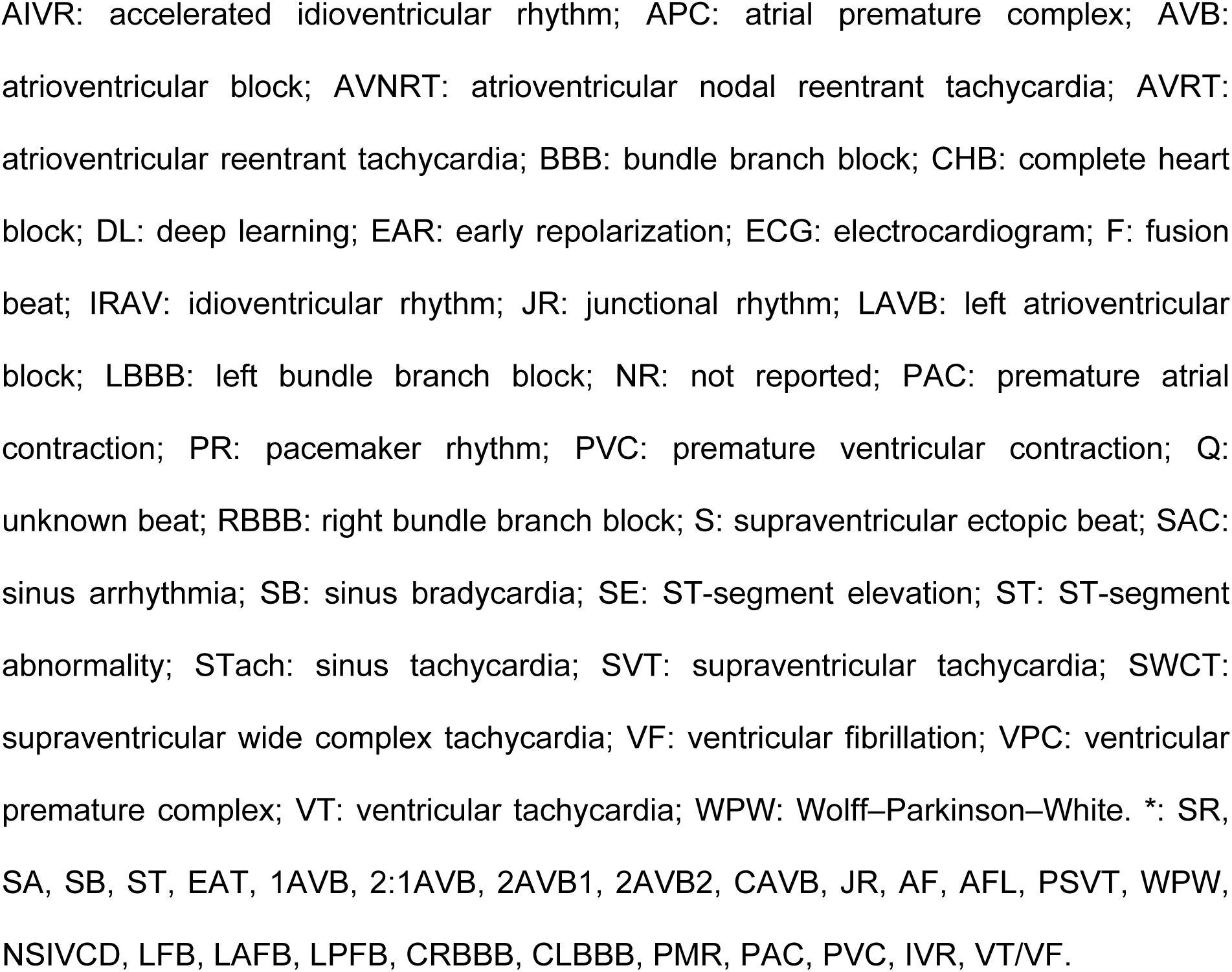
Baseline characteristics of the included studies.

| Study | Country | External validation | Number of ECGs in test set | Number of patients | Total number of ECGs | Mean Age years | Male (%) | Number of arrhythmias in test set (%) | Types of arrhythmias |
| --- | --- | --- | --- | --- | --- | --- | --- | --- | --- |
| Bridge 2022 | UK | No | 172 | 1172 | 1172 | 71.5 | 25.6 | 38 (22.1) | AF; AFL; PAVC; AVB; SVT; WPW; PR; JR |
| Cai 2020 | China | No | 3312 | 11994 | 16557 | 57 | 47.4 | 743 (22.4) | AF; Other |
| Chang 2021 | Canada | Yes | 116 | 38899 | 65932 | NR | NR | 107 (92.2) | AF; AFL; APB; VB; CHB; EAR; FRAV; PSVT; SAC; ST; VPB |
| Elsheiky 2025 | Egypt | No | 35139 | 32 | 175694 | 58 | 53 | 4429 (12.6) | S; V; F; Q |
| Gliner 2020 | Russia | No | 5830 | 6866 | 41830 | NR | 53.6 | 1007 (17.3) | AF; I-AVB; LBBB; RBBB; PAC; PVC; STD; ST; STE ST |
| Higuchi 2023 | USA | No | 218 | 725 | 1505 | NR | NR | 110 (50.4) | AVNRT; AVRT; AT |
| Jeong 2021 | China | No | 2954 | 6877 | 6877 | NR | 53.7 | 2560 (86.7) | AF; I-AVB; LBBB; RBBB; PAC; PVC; STD; ST; STE ST |
| Jekova 2022 | Bulgaria | No | 6978 | NR | 71545 | NR | NR | 559 (8) | AF; AFL |
| Jo 2020 | South Korea | Yes | 12914 | 62871 | 228364 | NR | NR | 1679 (13) | AF |
| Kashou 2022 | USA | Yes | 235 | 235 | 623 | 69.88 | NR | 235 (100) | SWCT; VT |
| Kolliyil 2025 | USA | Yes | 1.072 | NR | 7472 | NR | NR | 968 (90.3) | AF; IAVB; LBBB; RBBB; PAC; PVC; STD; STE |
| Li 2021 | China | No | 10521 | 51579 | 51579 | NR | NR | 2955 (28) | NSR; ST; SB; APC; VPC; AF; I-AVB |
| Li 2024 | China | No | 54 | 165 | 165 | 51.31 | 64.8 | 54 (1) | SVT; VT |
| Lin 2025 | Taiwan | Yes | 4609 | NR | 22130 | 70.4 | 60.4 | 869 (18.9) | 23 Classes * |
| Ribeiro 2020 | Brazil | No | 827 | 1676384 | 2322513 | 53.64 | 39.7 | 187 (22.6) | I-AVB; RBBB; LBBB; SB; AF; ST |
| Sadeghi 2024 | USA | No | 1126 | NR | 10344 | NR | 54 | 779 (69.2) | AF, I-AVB, LBBB, RBBB, SB, STach |
| Smith 2019 | USA | No | 1473 | NR | 24123 | NR | NR | 84 (5.7) | AFL; II-AVB; III-AVB; CAVB; RBBB; AT; SB; ST; PR; LBBB |
| Ye 2022 | China | No | 2954 | 6877 | 2954 | NR | 53.78 | 2560 (86.7) | AF; I-AVB; LBBB; RBBB; PAC; PVC; STD; STE |
| Zhu 2020 | China | Yes | 828 | 70692 | 180112 | 57.9 | 54.2 | NR | NSR; I-AVB; PR; VT; PVC; AF; AT; 2AVB-T1 |

Abbreviations: 1AVB, 2AVB-I, 2AVB-II: first- and second-degree atrioventricular blocks (Mobitz I and II); AFL: atrial flutter; AF: atrial fibrillation; AI: atrial impulse anomaly;
AIVR: accelerated idioventricular rhythm; APC: atrial premature complex; AVB: atrioventricular block; AVNRT: atrioventricular nodal reentrant tachycardia; AVRT: atrioventricular reentrant tachycardia; BBB: bundle branch block; CHB: complete heart block; DL: deep learning; EAR: early repolarization; ECG: electrocardiogram; F: fusion beat; IRAV: idioventricular rhythm; JR: junctional rhythm; LAVB: left atrioventricular block; LBBB: left bundle branch block; NR: not reported; PAC: premature atrial contraction; PR: pacemaker rhythm; PVC: premature ventricular contraction; Q: unknown beat; RBBB: right bundle branch block; S: supraventricular ectopic beat; SAC: sinus arrhythmia; SB: sinus bradycardia; SE: ST-segment elevation; ST: ST-segment abnormality; STach: sinus tachycardia; SVT: supraventricular tachycardia; SWCT: supraventricular wide complex tachycardia; VF: ventricular fibrillation; VPC: ventricular premature complex; VT: ventricular tachycardia; WPW: Wolff–Parkinson–White. \*: SR, SA, SB, ST, EAT, 1AVB, 2:1AVB, 2AVB1, 2AVB2, CAVB, JR, AF, AFL, PSVT, WPW, NSIVCD, LFB, LAFB, LPFB, CRBBB, CLBBB, PMR, PAC, PVC, IVR, VT/VF.

### Artificial Intelligence Models

**Table 2** illustrates the variety of AI approaches and the diagnostic references adopted. Most studies employed convolutional neural networks (CNNs), with 11 of the 20 investigations using these architectures, including Inception-V3, PyTorch, and ECG-oriented convolutional networks. One study used a deep densely connected neural network. Three studies applied traditional machine-learning classifiers, such as logistic regression and gradient-boosting approaches, while three additional investigations implemented hybrid pipelines that combined long short-term memory (LSTM) architectures with CNN, CNN-based feature extractors with XGBoost classifiers, or feature-engineering stages with generic deep-learning models. One investigation relied on a pure LSTM architecture, and another adopted an explainable deep learning (XDL) structure. The diagnostic reference standards also varied, encompassing cardiologists, electrophysiologists, or consensus panels, depending on the study design.

**Table 2:** Characteristics of AI models.

| Study | AI Algorithm | AI sensitivity % | AI specificity % | AI AUC | Accuracy % | Reference Standard |
| --- | --- | --- | --- | --- | --- | --- |
| Bridge 2022 | CNN (InceptionV3) | 81.6 | 99.3 | 93.2 | NR | Electrophysiologist |
| Cai 2020 | CNN (DDNN) | 99.19 | 99.44 | 99.90 | 99.35 | Cardiologists |
| Chang 2020 | LSTM | 84 | NR | NR | 90 | Cardiologists |
| Elsheiky 2025 | CNN | 99.48 | 99.87 | NR | 99.48 | NR |
| Gliner 2020 | CNN (Pytorch Framework with Python) | NR | NR | NR | 96 | Cardiologists |
| Higuchi 2023 | CNN | 81 | 94.5 | 93.4 | NR | Electrophysiologists |
| Jeong 2021 | CNN | 78 | 96 | 87 | NR | NR |
| Jekova 2022 | DNN (DenseNet) | 91.8 | 95.8 | NR | NR | NR |
| Jo 2020 | XDL | 98.5 | 99.3 | 99.7 | 99.2 | Cardiologists |
| Kashou 2022 | ML (Logistic regression) | 85.5 | 91.5 | 95 | 88.5 | Cardiologists and non-cardiologists |
| Kolliyil 2025 | ML (XGBoost) | 96.9 | 58.7 | 92 | 68 | ECG labels by cardiologists |
| Li 2021 | CNN (Inception-V3) | 95.43 | 96.75 | NR | 98.46 | Experts and cardiologists |
| Li 2024 | ML (GBM) | 96.97 | 90.48 | 97 | 94.44 | NR |
| Lin 2025 | CNN (ECG12Net) | 95.4 | 98.7 | 98.92 | NR | Cardiologists |
| Ribeiro 2020 | CNN | 92.5 | 98.6 | NR | NR | NR |
| Sadeghi 2024 | LSTM plus CNN | NR | NR | 81.9 | 73.3 | NR |
| Smith 2019 | CNN (Cardiologs and Veritas) | 92.5 | 99.4 | NR | 99.2 | Emergentist and cardiologist |
| Ye 2022 | CNN (CBI-DF-XGBoost) | 81.4 | NR | 89.8 | 96.8 | NR |
| Zhu 2020 | CNN | 86.7 | 99.5 | 98.3 | NR | Cardiologists |
| Zvuloni 2023 | FE plus DL (Feature engineering combined with Deep learning) | NR | NR | 90 | NR | Electrophysiologists |

Abbreviations: CNN = convolutional neural network; DNN = deep neural network; DDNN = deep densely connected neural network; LSTM = long short-term memory; ML = machine learning; XDL = explainable deep learning; GBM = gradient boosting machine; DL = deep learning; XGBoost = extreme gradient boosting; AUC = area under the receiver operating characteristic curve; NR = not reported.

### Pooled Metrics

The overall pooled analysis demonstrated a sensitivity of 94.0% (95% CI 90.8–96.2; I² = 96.9%) (**Fig. 2**) and a specificity of 98.7% (95% CI 97.3–99.3; I² = 98.3%) (**Fig. 3**), in the context of an overall arrhythmia prevalence of 21.7%. The pooled area under the curve (AUC) was 0.982 (95% CI 0.965–0.986) (**Fig. 4**).

**Fig. 2.**
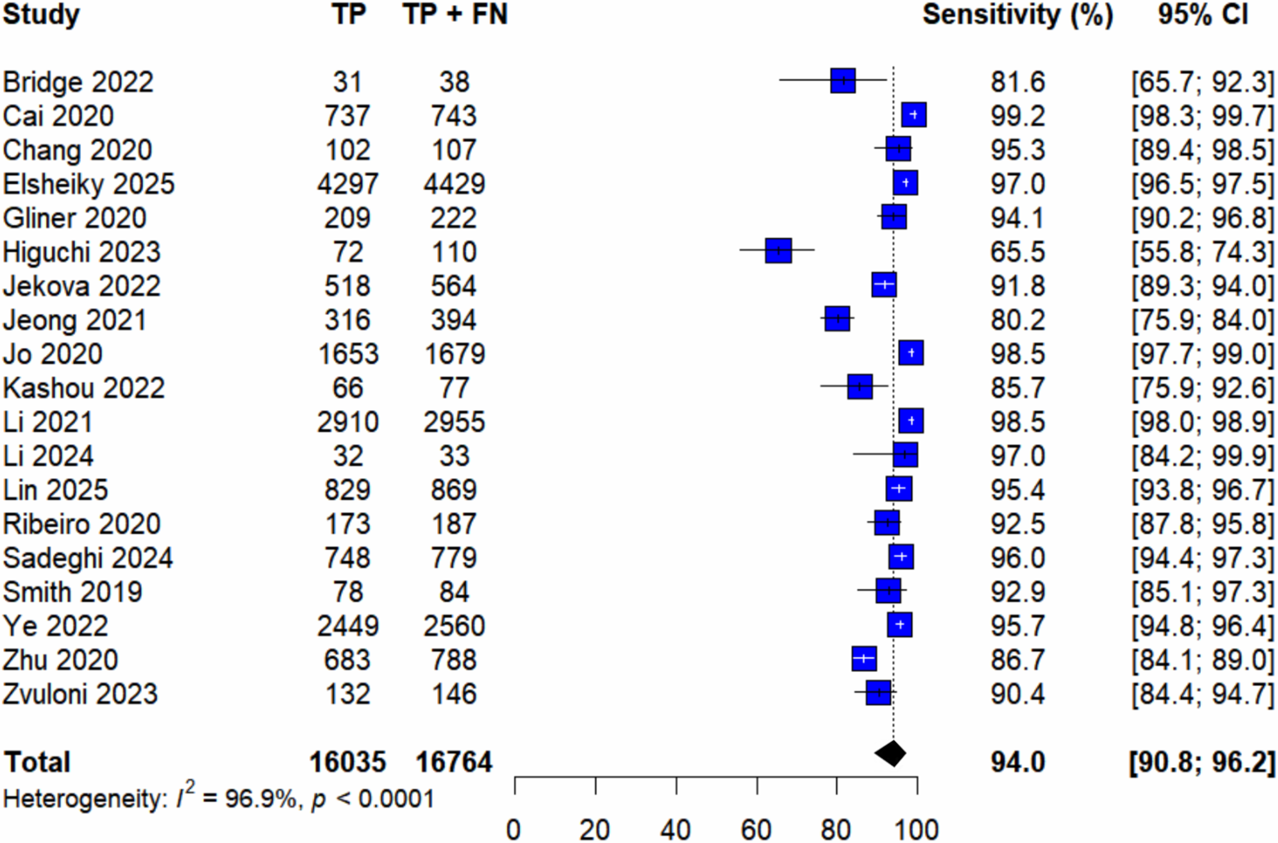
Forest plot for pooled model sensitivity. TP = True positives; FN = False Negatives.

**Fig. 3.**
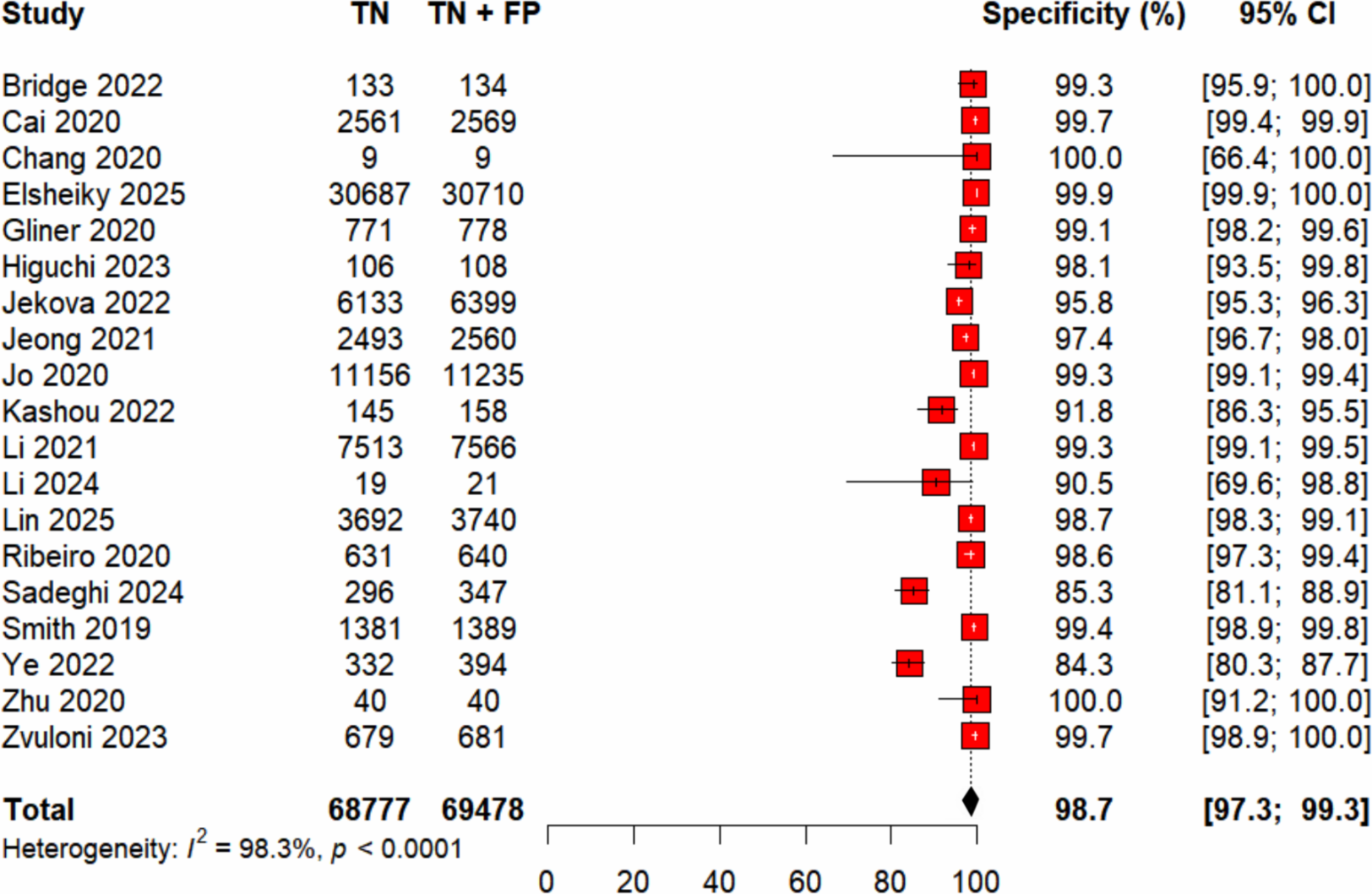
Forest plot for pooled model specificity. TN = True Negatives; FP = False Positives.

**Fig. 4.**
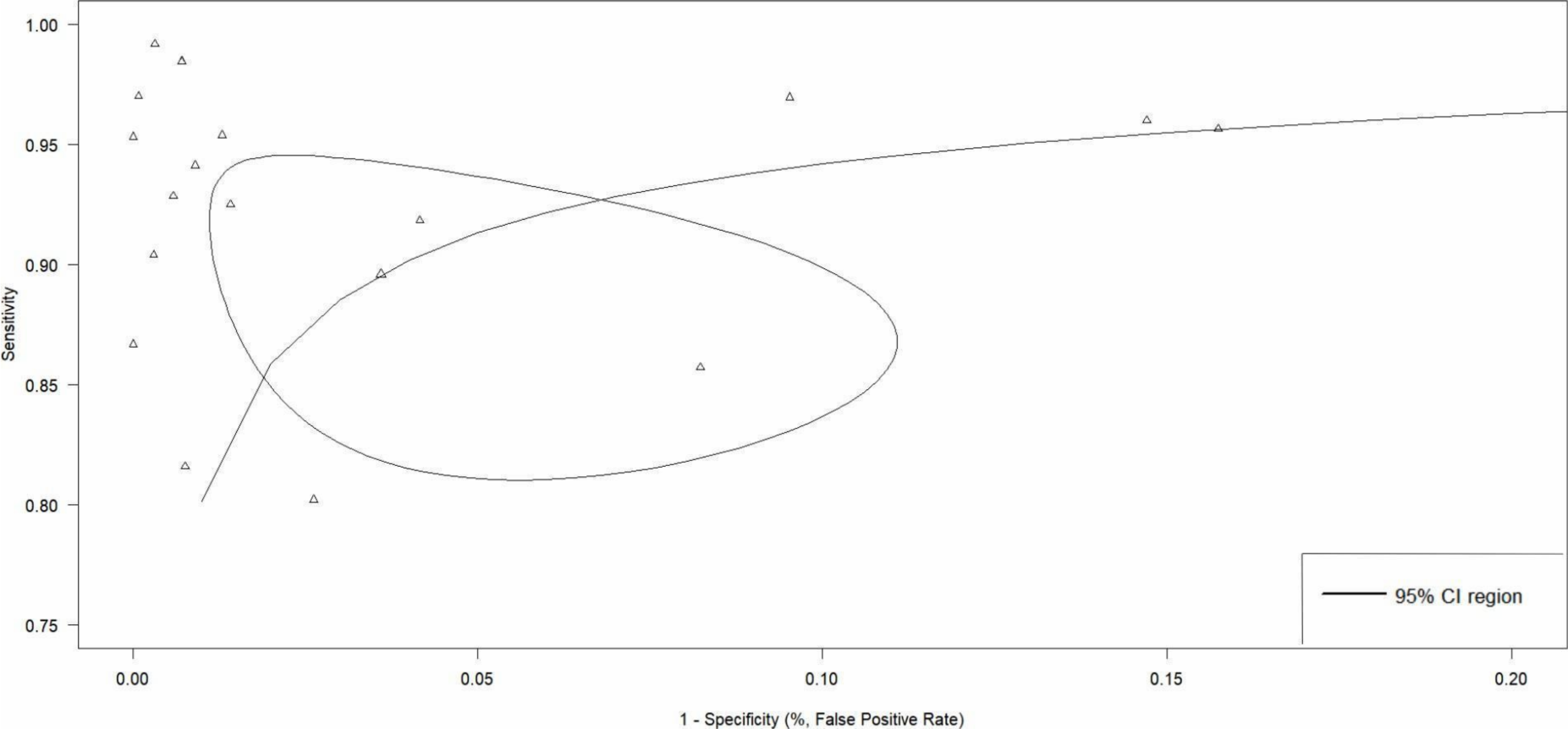
SROC curve for AI diagnosis of arrhythmia on ECG*. *Scales were adjusted for readability

### Subgroup Analyses

A pooled analysis focused specifically on AF detection **(S1, S2),** demonstrated a sensitivity of 92.6% (95% CI 86.4–96.1; I² = 96.0%) and a specificity of 99.1% (95% CI 98.4–99.5; I² = 95.9%), in the context of an overall AF prevalence of 10.7%. The AUC for AF discrimination was 0.988 (95% CI 0.981–0.989). A second subgroup analysis restricted to CNN-based architectures **(Figs. S3, S4)**, yielded a sensitivity of 93.9% (95% CI 89.2–96.6; I² = 97.6%) and a specificity of 98.7% (95% CI 96.9–99.5; I² = 98.5%), in the context of an overall arrhythmia prevalence of 22.4%. The corresponding AUC was 0.982 (95% CI 0.959–0.987). For the external validation subanalysis (n = 6 studies) **(Figs. S5, S6)**, the pooled sensitivity was 96.9% (95% CI 89.2–99.1; I² = 98.4%) and specificity 95.6% (95% CI 77.6–99.3; I² = 99.2%), with an AUC of 0.983 (95% CI 0.873–0.989), with an arrhythmia prevalence of 13.3%. The analysis of studies with a similar prevalence of abnormal classes **(Figs. S7, S8)** demonstrated comparable performance but with moderately lower heterogeneity, including sensitivity of 94.5% (95% CI 88.9–97.14; I² = 87.8%) and specificity of 99.3% (95% CI 98.7–99.6; I² = 72.5%), with an AUC of 0.989 and an overall arrhythmia prevalence of 24.3%.

### Meta-regression

Based on the methodological characteristics of the included studies, we examined four prespecified factors—ECG dataset size, task type (single-versus multi-arrhythmia classification), arrhythmia prevalence, and AI model architecture—using meta-regression **(Figs. S9, S10, S11, S12)**. The analyses showed that none of these covariates produced statistically significant modifications in sensitivity or specificity. Likewise, the patterns observed across the four models did not suggest substantial reductions in between-study variability. These findings indicate that differences in dataset magnitude, diagnostic complexity, prevalence distribution, or model architecture do not fully account for the substantial heterogeneity observed in the meta-analysis.

### Quality Assessment, Publication Bias, and Certainty of Evidence

Using the QUADAS-2 tool, overall risk of bias was predominantly low across the four domains (**Fig. 5**). In the Patient Selection domain, most studies used large and representative ECG datasets with clearly defined inclusion criteria, minimizing selection bias, although a few presented unclear reporting. The Index Test domain was also considered predominantly low risk, as AI models were applied in a standardized manner, generally blinded to reference results. The Reference Standard domain showed low risk in most studies because the ground truth was consistently established by cardiologists or validated clinical annotations, with only a few cases of unclear reporting. Similarly, the Flow and Timing domain presented uniformly low risk, given the absence of inappropriate exclusions or significant delays between index testing and reference assessment.

**Fig. 5.**
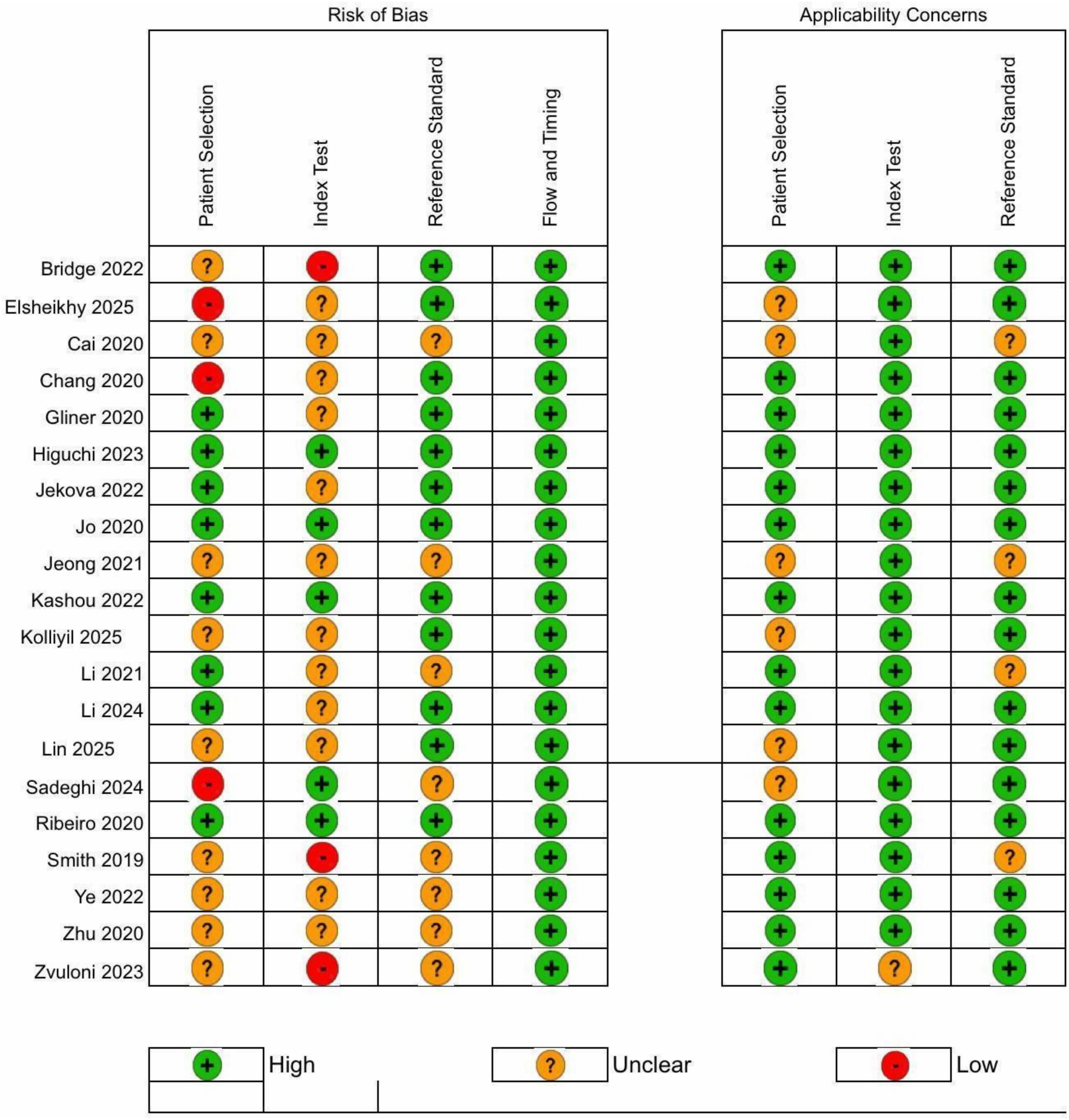
Risk of bias assessment by QUADAS-2 tool.

Regarding applicability concerns, all three domains, Patient Selection, Index Test, and Reference Standard, were judged as low concern overall, since the populations, diagnostic targets, and evaluation methods were aligned with the research question. Only a few studies showed minor uncertainty related to the representativeness of retrospective datasets.

The included studies demonstrated a predominantly low risk of bias and low applicability concerns, supporting the robustness of the pooled diagnostic estimates.

The funnel plot **(Fig. S13)** revealed a symmetrical distribution of studies around the pooled effect estimate, with points spread evenly across both sides, suggesting no small-study effects. Consistently, the Egger regression test showed no significant asymmetry (t = 0.47; df = 17; p = 0.6438; bias = 1.5380; SE = 3.2669), indicating no evidence of publication bias.

According to the GRADE framework **(Table S1)**, the overall certainty of the evidence was rated as high, supported by the predominance of low or unclear risk of bias in most domains, robust pooled estimates for both specificity and sensitivity, and the absence of publication bias.

## Discussion

To the best of our knowledge, this is the largest meta-analysis evaluating AI models for arrhythmia detection among studies restricted to 12-lead ECGs, including twenty studies encompassing more than 5.5 million ECGs in total and 92,159 in testing cohorts. Our results demonstrated high diagnostic performance in detecting cardiac arrhythmias, with a pooled sensitivity of 94.0% (95% CI 90.8–96.2), specificity of 98.7% (95% CI 97.3–99.3), and an AUC of 0.982 (95% CI 0.965–0.986). Most studies employed CNNs or related neural network architectures. Subgroup analyses demonstrated consistent performance when specifically assessing discrimination of AF, as well as studies utilizing CNN-based architectures.

Our findings are consistent with previous investigations demonstrating that deep learning models can achieve high diagnostic accuracy across multiple arrhythmia types. The literature has repeatedly shown that both single-lead and 12-lead ECG can reach specialist-level performance, reinforcing the translational potential of AI-assisted ECG interpretation ^7,13,25^. In this context, the strong pooled metrics observed in our analysis demonstrate that these models perform robustly across varying datasets and methodological approaches. Xie et al. (2024) recently conducted a meta-analysis evaluating AI models for ECG-based AF detection without restricting the number of leads, reporting a sensitivity and specificity of 97%. Our analysis encompassed a broader arrhythmia spectrum and a substantially larger volume of 12-lead ECGs, providing a more comprehensive evaluation of AI performance across diverse rhythms and study designs, while better reflecting practical applicability given the central role of the 12-lead ECG in routine practice ^34^. This high effectiveness can translate into improved diagnostic precision, earlier detection of asymptomatic or subclinical arrhythmias, reduced interpretation time, and enhanced support for clinical decision-making ^16,27^. Taken together, these data support the potential role of AI-enabled ECG analysis as a useful tool for arrhythmia classification.

AI architecture may influence the efficacy of ECG-based arrhythmia classification. CNNs are the most commonly used for arrhythmia classification because they perform hierarchical feature extraction without the need for manual engineering, enabling robust discrimination of subtle waveform variations ^31,35,36^. In our study, CNN-based models were the most common architecture utilized, and performed well for overall arrhythmia classification (sensitivity of 93.9%; specificity of 98.7%; AUC of 0.982).

Our findings also support the specific role of ECG-based AI for the detection of AF, a particularly common and morbid arrhythmia. Among studies that analyzed AF detection, specificity was nearly perfect, along with sensitivity >90%. The strong performance observed in our AF subgroup analyses aligns with evidence that AI-enabled ECG analysis can identify electrophysiologic signatures associated with AF even in sinus rhythm, suggesting that deep learning models capture subtle risk markers beyond human visual interpretation ^37^. The ability to detect AF accurately is particularly relevant for efforts to reduce AF-related stroke risk and enable mass AF screening strategies ^38^.

Our findings highlight a critical need for greater standardization in the field of ECG-based AI. Despite consistently high sensitivity and specificity, significant heterogeneity was observed in our meta-analysis, and our subgroup analyses – including those based on model type, arrhythmia-specific studies, external validation – and meta-regression were unable to fully explain its origin. Of the secondary analyses performed, only our subgroup analysis of studies with a similar prevalence of abnormal classes resulted in moderately lower heterogeneity, suggesting differences in class balance (i.e., patient characteristics) as one driver. Additional heterogeneity may stem from a variety of factors, including differences in AI architectures, dataset size, population characteristics, and the number of arrhythmia classes evaluated, which ranged from single-rhythm models to multiclass frameworks involving up to 23 arrhythmias ^27^. Variations in reference standards, ECG preprocessing methods, and reporting metrics may further contribute to heterogeneity. On balance, our results compel the need for data annotation and harmonized methodological protocols to promote reproducibility across centers ^39^, as well as more consistent reporting practices to enhance comparability between studies. There are recent guidelines for reporting AI research, such as the EHRA scientific statement and the AHA TRUE-AIM report, which could help to mitigate these variations ^40,41^. Importantly, although statistical heterogeneity was high, all individual study effects were consistently favorable, indicating that the variability observed did not meaningfully alter the clinical interpretation of our pooled estimates.

Despite optimal pooled performance, several authors have emphasized that real-world applicability requires validation in larger and more diverse populations ^7^. For example, the EAGLE and BEAGLE trials demonstrated improved detection of systolic dysfunction and AF using AI-enhanced ECG ^42,43^. Although AI achieves high accuracy for arrhythmia detection from 12-lead ECGs, external validation across independent cohorts remains limited ^44^. For instance, the EHRA scientific statement reported that only 58% of AI studies in AF management included external testing, highlighting a gap that restricts clinical translation ^41^. In our external validation subanalysis, a slightly better sensitivity and worse specificity were found, indicating that AI models can still perform well when evaluated in external settings. Lower specificity may stem from the distributional shift between real-world clinical use and model development ^45^. Overall, however, the relative dearth of out-of-sample validation underscores a persistent gap in the field and compels future studies to prioritize external assessment to ensure generalizability in support of eventual clinical deployment in broad and varied settings.

### Limitations

Several limitations must be acknowledged. First, the high heterogeneity observed across studies represents a significant limitation, although it did not materially alter the clinical interpretation of our pooled results. As noted above, heterogeneity likely reflects methodological and population differences and underscores the need for greater standardization in future ECG-AI research. Second, variations in data availability, given that some studies reported patient-level metrics while others provided only aggregate measures, constrained the depth of potential subgroup analyses. Third, although the QUADAS-2 assessment indicated that most studies presented a low risk of bias, several remained at high or unclear risk in some domains. Fourth, our data extraction was limited by the information available in the published literature, and several studies had to be excluded because the metrics required to derive diagnostic accuracy estimates could not be calculated from the reported results. Finally, the predominance of retrospective designs limits data representativeness and increases the likelihood of overlap between development and validation datasets, particularly when public repositories were reused across publications. This also restricts clinical validation and raises the possibility of overfitting, especially for models trained and tested within single-institution or limited datasets.

## Conclusion

In a large meta-analysis including 20 studies and over 5 million ECGs, AI models applied to 12-lead ECGs demonstrate excellent accuracy for arrhythmia detection. Pooled metrics support diagnostic utility and provide a basis for potential integration into clinical workflows. Due to substantial heterogeneity across studies, however, future work is needed to promote standardization of study design, reporting, and external validation frameworks.

## Data Availability

All data used in this study are derived from published articles and publicly available sources, which are cited within the manuscript and listed in the references.

