## Supplementary material for "Diagnostic Accuracy of Artificial Intelligence for Arrhythmia Detection Using the 12-Lead Electrocardiogram: A Systematic Review and Meta-Analysis"

Figure **S1** – Forest plot for Atrial Fibrillation Sensitivity

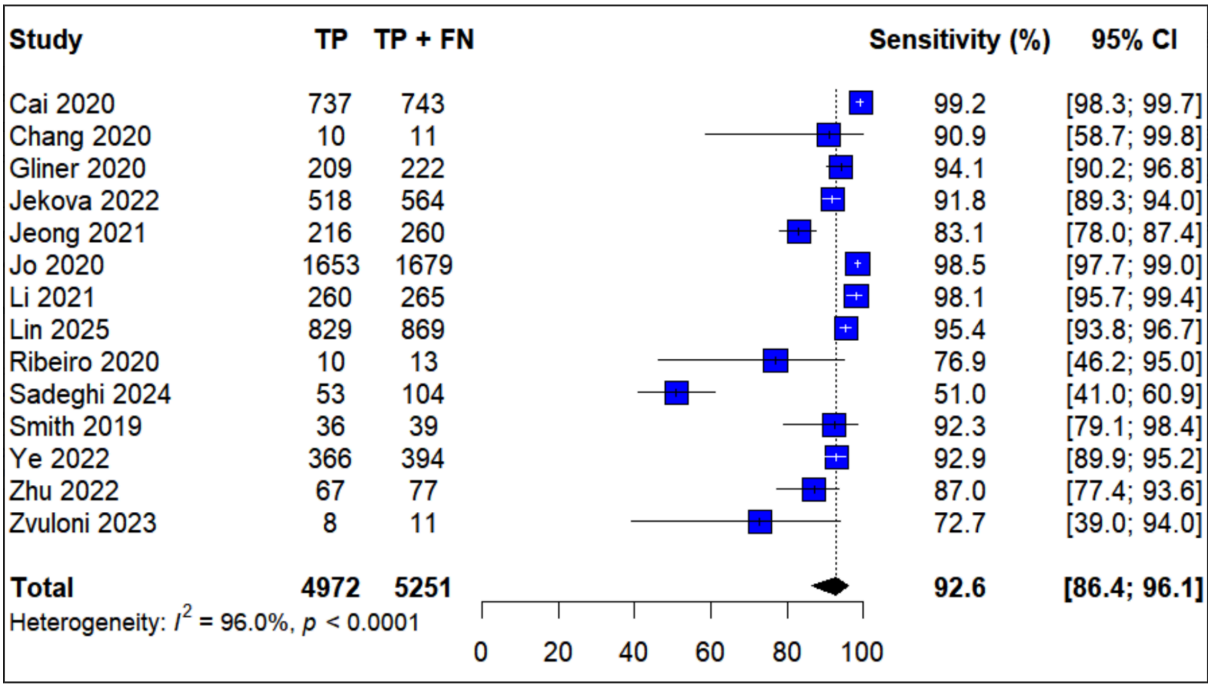

Figure **S2** – Forest plot for Atrial Fibrillation Specificity

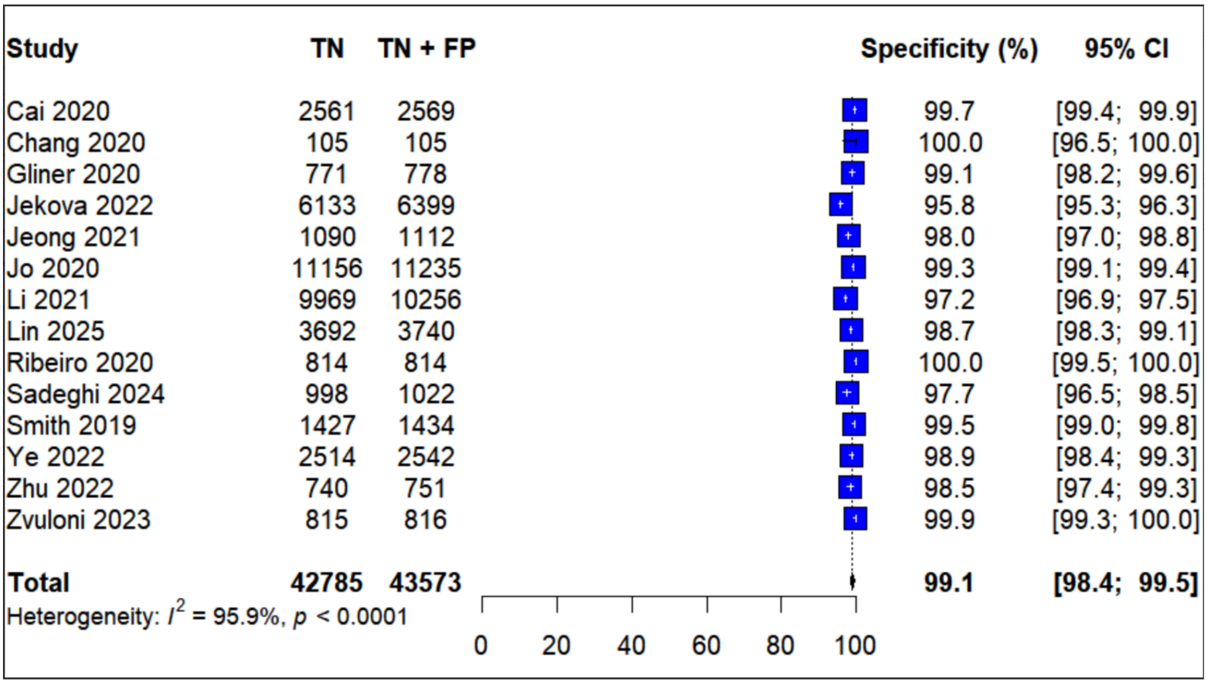

Figure **S3** – Forest plot for CNN models Sensitivity

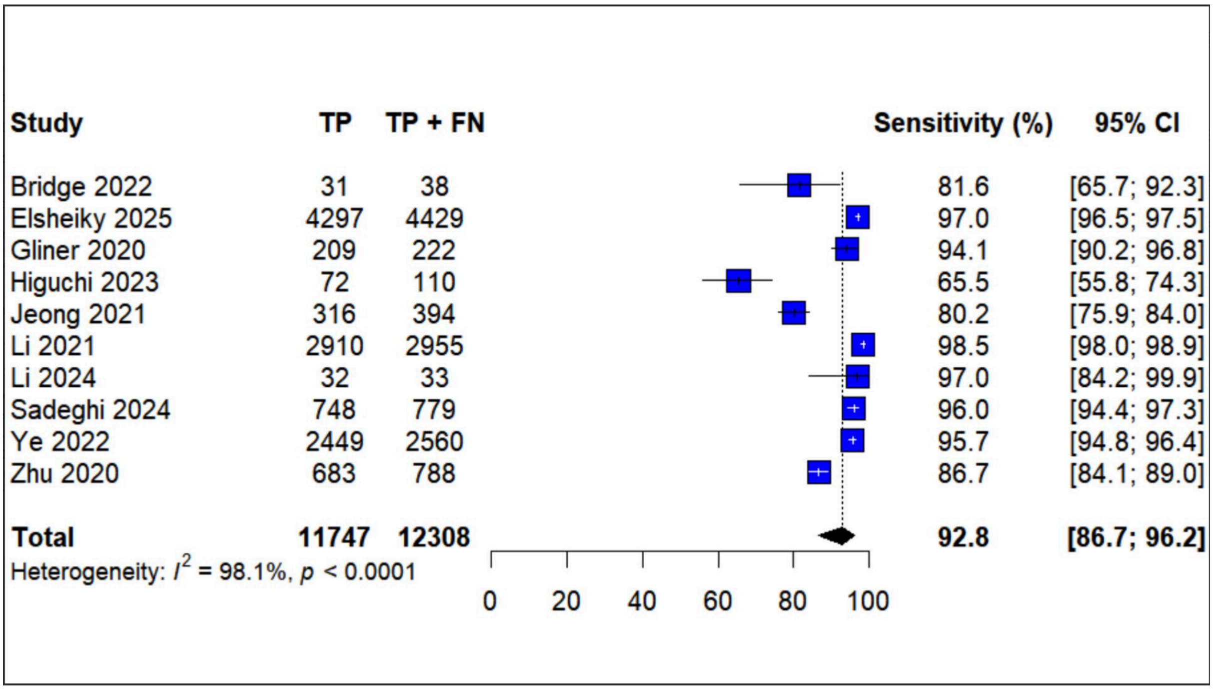

Figure **S4** – Forest plot for CNN models Specificity

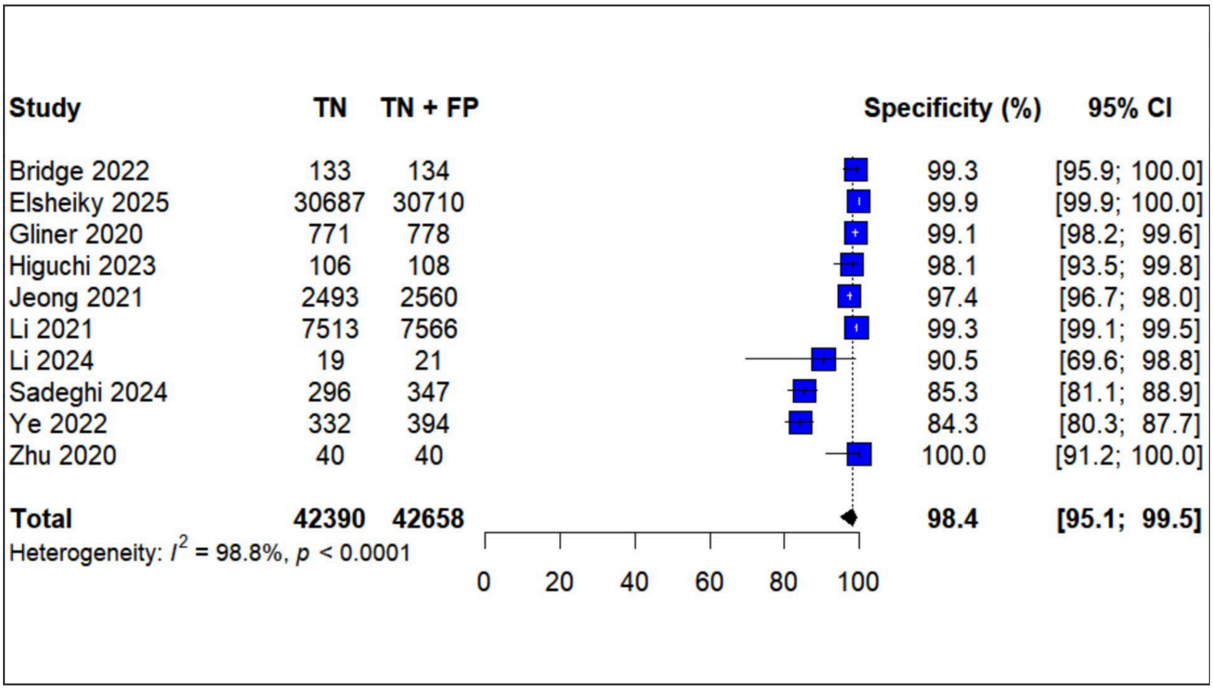

Figure **S5** – Forest plot for External validation Sensitivity

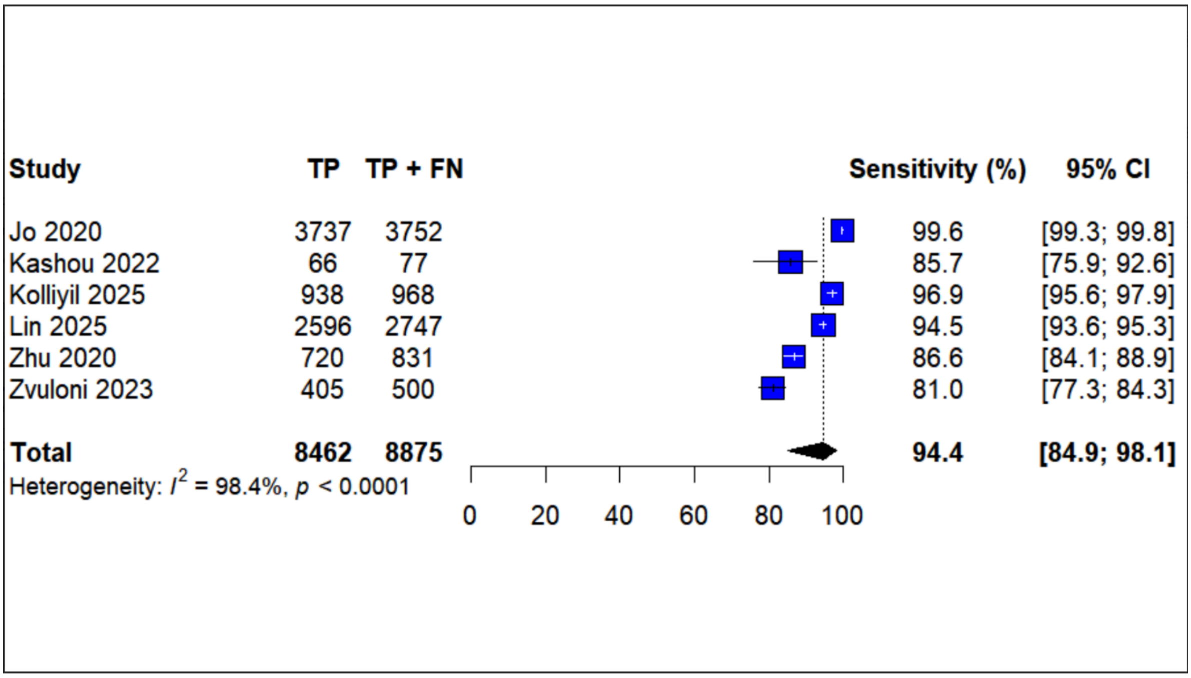

Figure **S6** – Forest plot for External validation Specificity

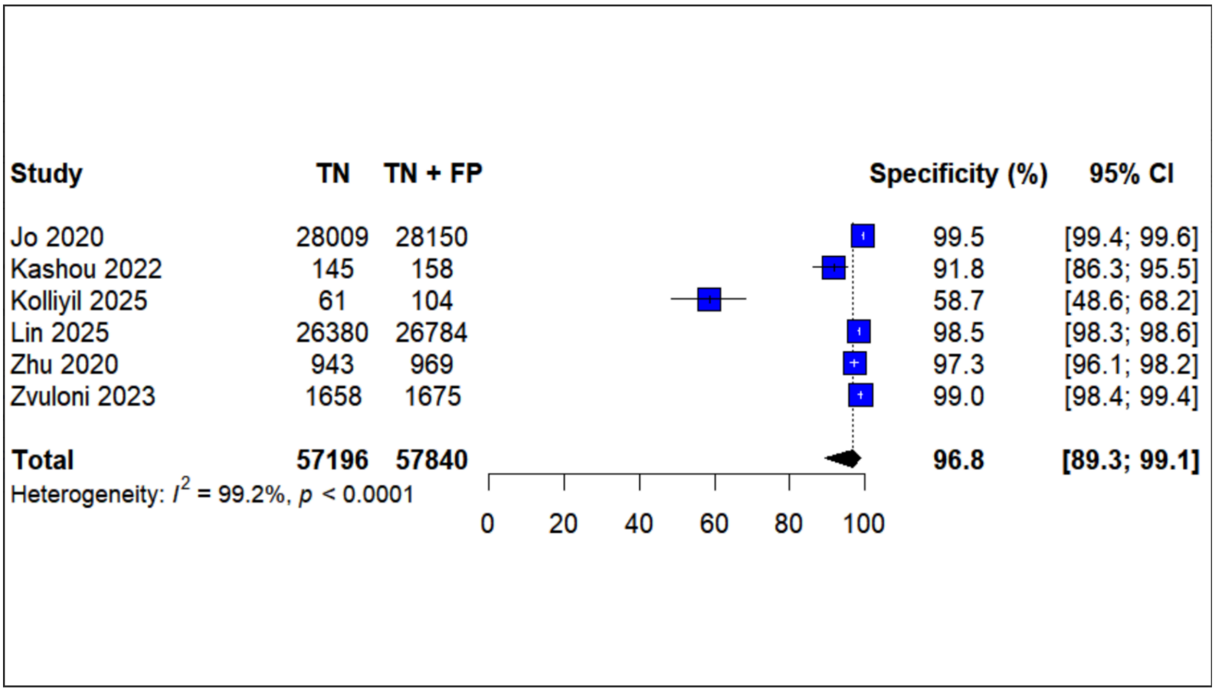

Figure **S7**– Forest plot for studies with similar arrhythmia prevalence Specificity

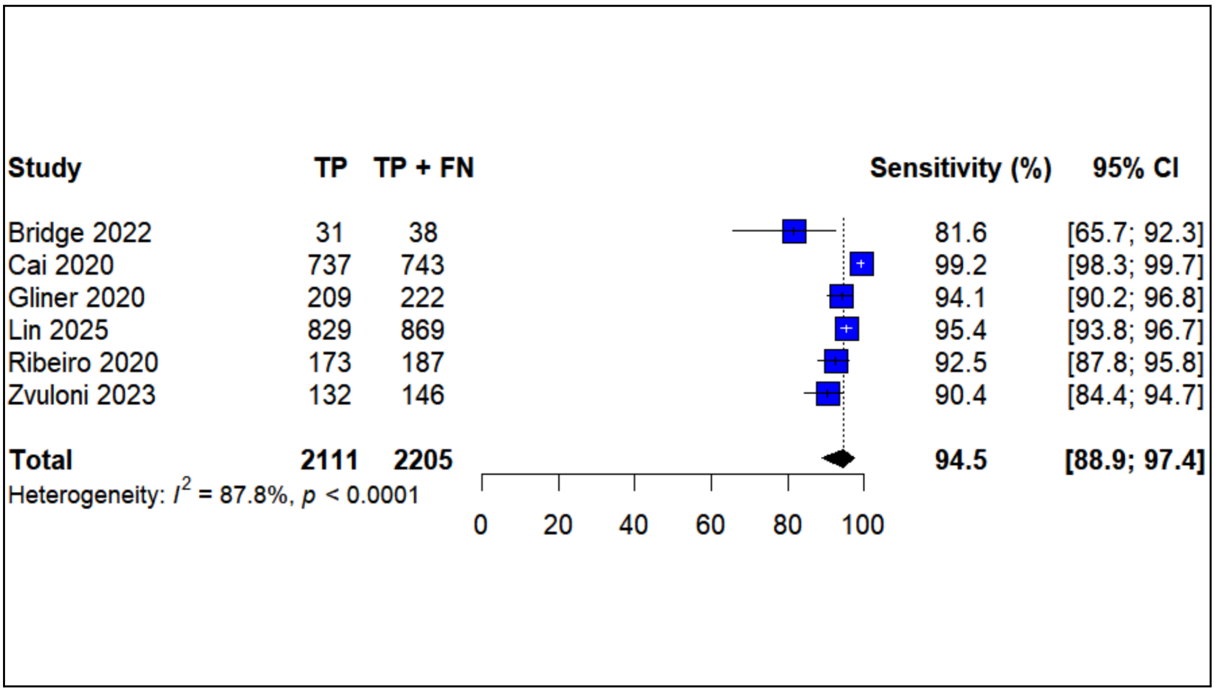

Figure **S8** – Forest plot for studies with similar arrhythmia prevalence Specificity

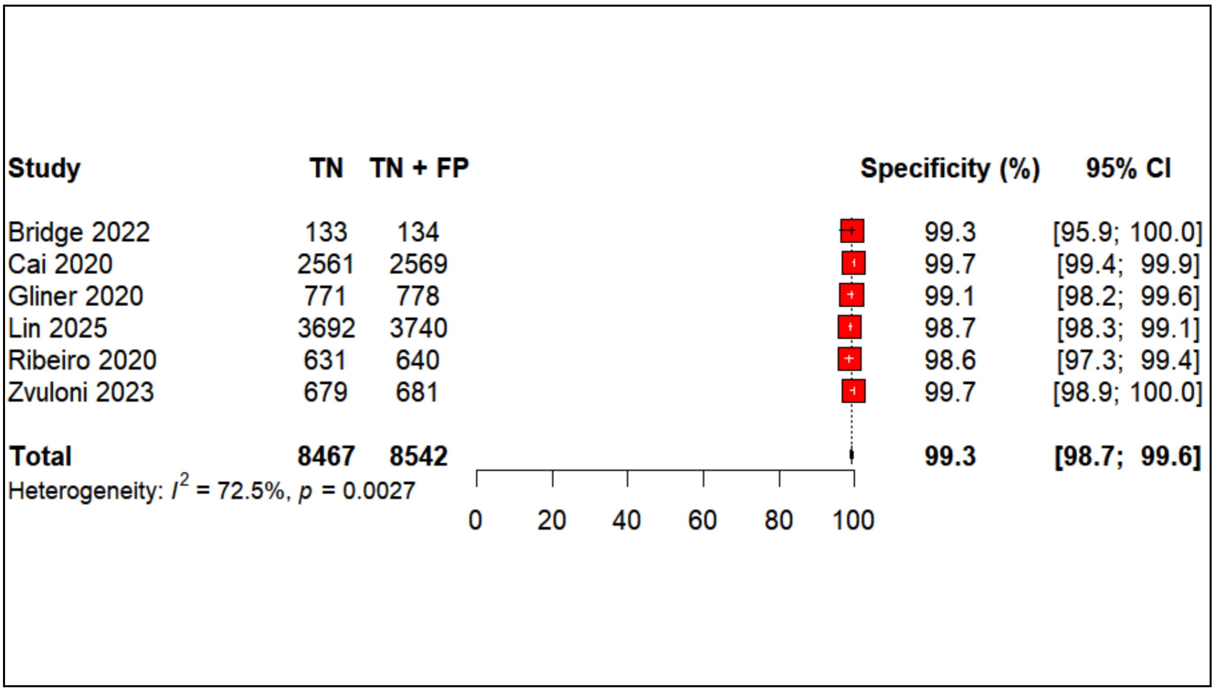

Figure **S9** – Meta-regression by AI model type

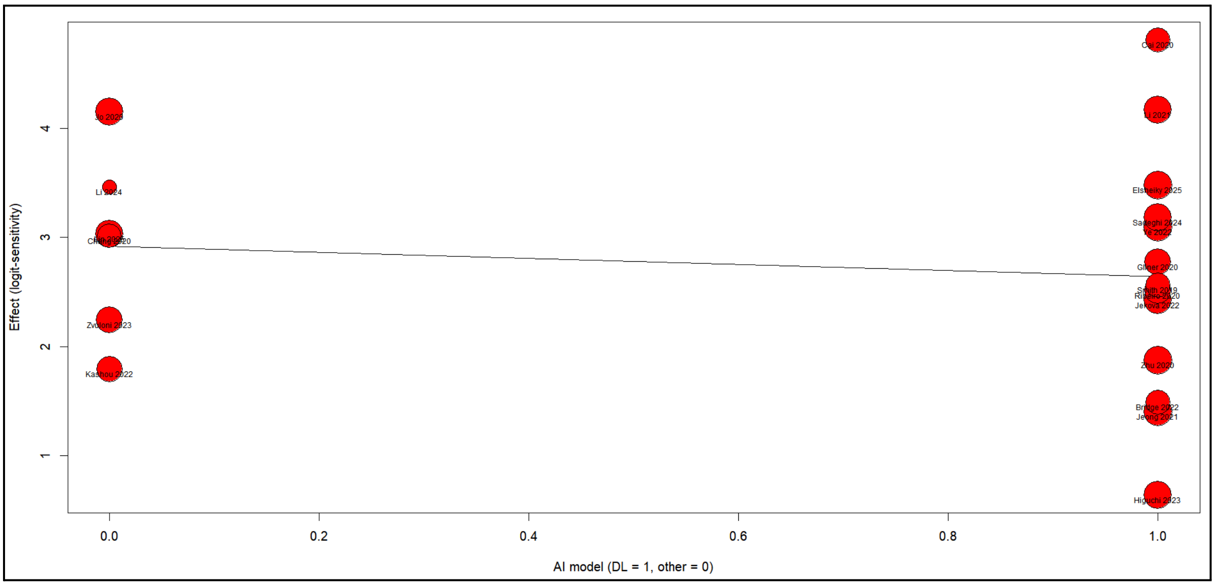

Figure **S10** – Meta-regression by classification strategy (binary vs multiclass)

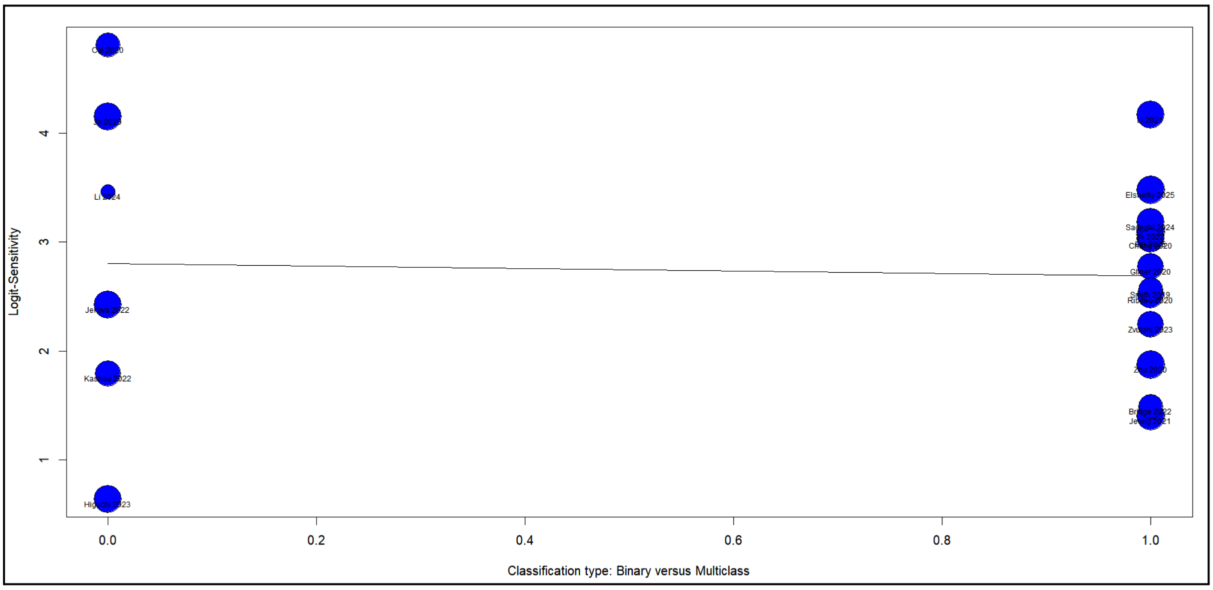

Figure **S11** – Meta-regression by ECG dataset size

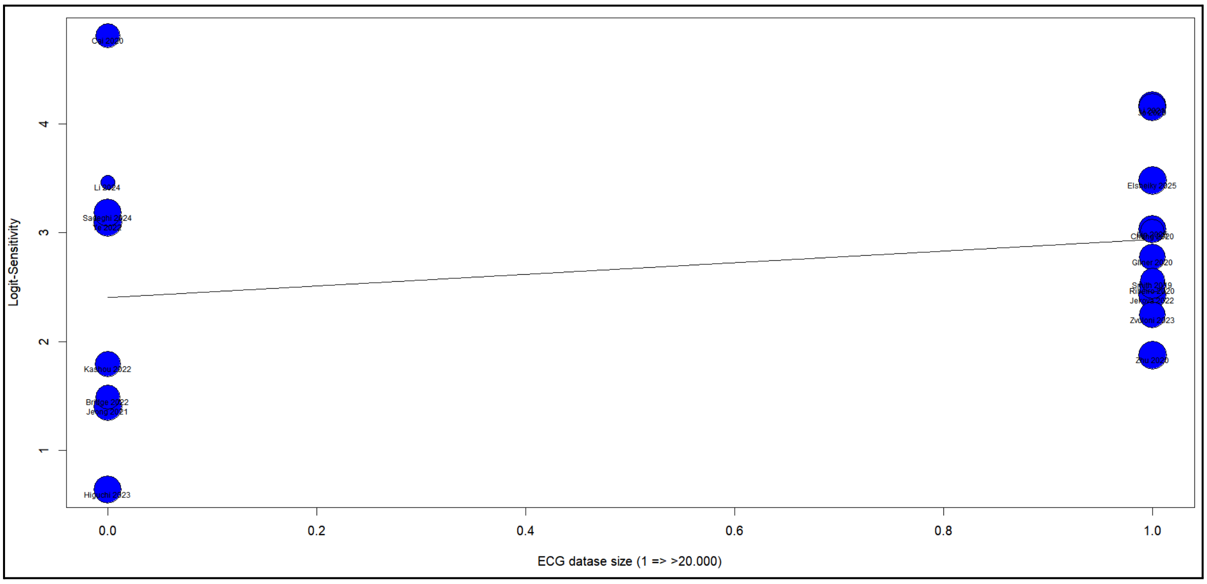

Figure **S12** – Meta-regression by arrhythmia prevalence

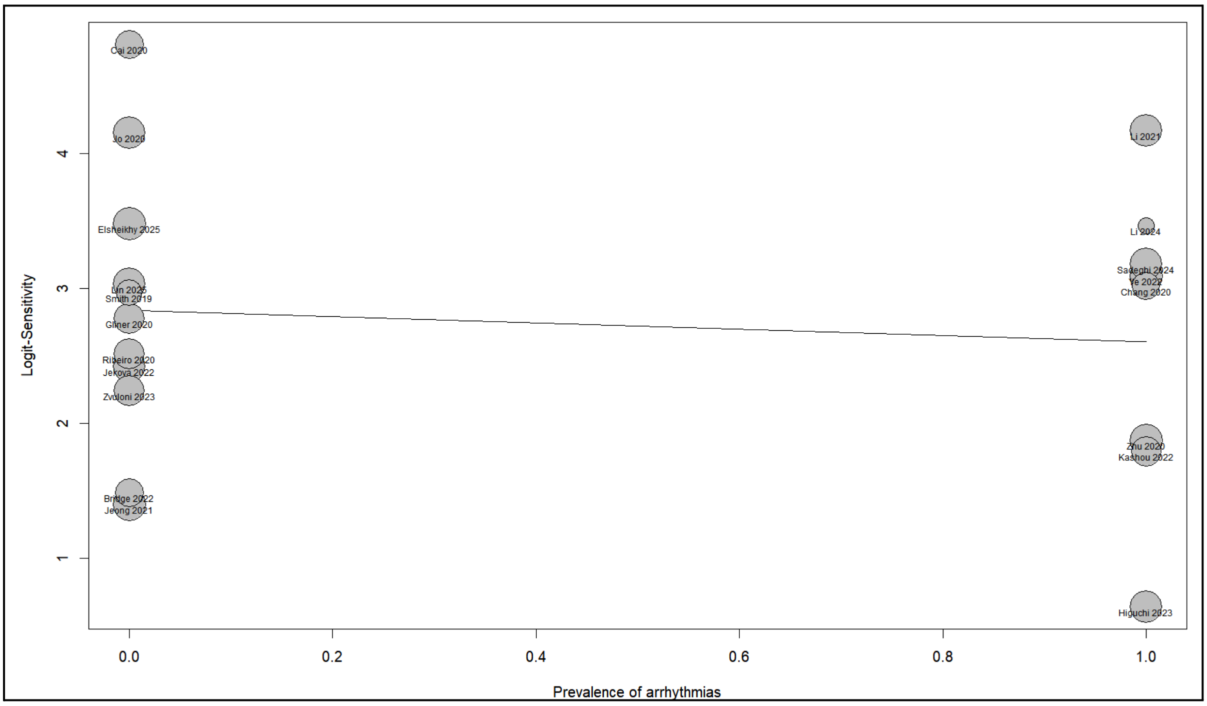

Figure **S13** – Funnel plot for publication bias

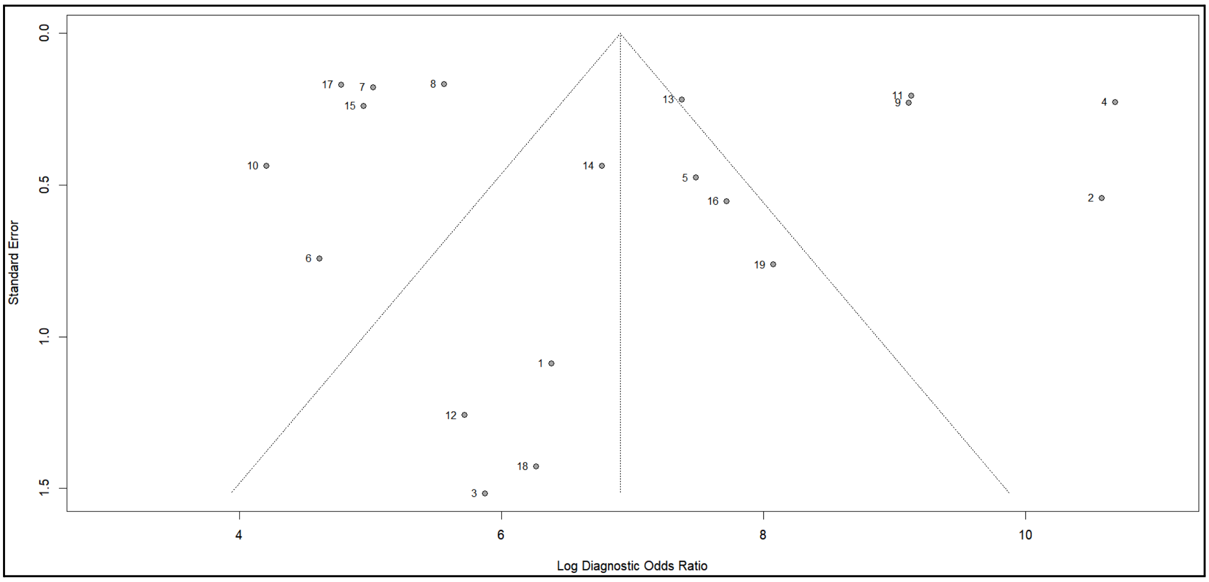

Table **S1** – GRADE assessment tool

| | |
|---|---|
| Sensitivity | 0.94 (95% CI: 0.91 to 0.96) |
| Specificity | 0.99 (95% CI: 0.97 to 0.99) |

| Outcome | № of studies (№ of patients) | Study design | Factors that may decrease certainty of evidence | | | | | Effect per 1.000 patients tested | Test accuracy CoE |
|---|---|---|---|---|---|---|---|---|---|
| | | | Risk of bias | Indirectness | Inconsistency | Imprecision | Publication bias | pre-test probability of 20% | |
| **True positives** (patients with arrhythmias) | 19 studies 16764 patients | cross-sectional (cohort type accuracy study) | not serious | not serious | not serious | not serious | strong association | 188 (182 to 192) | ⊕⊕⊕⊕ High |
| **False negatives** (patients incorrectly classified as not having arrhythmias) | | | | | | | | 12 (8 to 18) | |
| **True negatives** (patients without arrhythmias) | 19 studies 69478 patients | cross-sectional (cohort type accuracy study) | not serious | not serious | not serious | not serious | strong association | 790 (778 to 794) | ⊕⊕⊕⊕ High |
| **False positives** (patients incorrectly classified as having arrhythmias) | | | | | | | | 10 (6 to 22) | |
